# Economic evaluation of *Wolbachia* deployment in Colombia: A modeling study

**DOI:** 10.1101/2024.07.01.24309774

**Authors:** Donald S. Shepard, Samantha R. Lee, Yara A. Halasa-Rappel, Carlos Willian Rincon Perez, Arturo Harker Roa

**Affiliations:** Heller School for Social Policy and Management, Brandeis University, Waltham, Massachusetts, USA; School of Government, University of Los Andes, Bogotá, Colombia

**Keywords:** Dengue, Colombia, *Wolbachia*, Cost-effectiveness analysis, Benefit-cost analysis, Mosquito control

## Abstract

**Background and Aims:** *Wolbachia* are bacteria that inhibit dengue virus replication within the mosquito. A cluster-randomized trial found *Wolbachia* reduced virologically-confirmed dengue cases by 77% and previous models predicted *Wolbachia* to be highly cost-effective in Indonesia, Vietnam, and Brazil. in Colombia, *Wolbachia* was introduced in the Aburrá Valley in 2015 and Cali in 2020. To inform decisions about future extensions, we performed economic evaluations of the potential expansion of *Wolbachia* deployments to 11 target Colombian cities.

**Methods:** We assembled quantities and the distribution by severity of reported dengue cases from Colombia’s national disease surveillance system and the health service provision registry (RIPS). An epidemiological panel of three experts estimated the shares of non-medical cases and adjustments for under-reporting and misclassifications. We determined costs (in 2020 US dollars) of treating dengue illness from the benchmark insurance tariff, RIPS data on treatment services per symptomatic dengue case, and the national government database for establishing insurance premiums. A cluster randomized trial quantified the effectiveness of *Wolbachia* against symptomatic dengue cases.

**Results:** Projecting impact over 10 years for Cali, we estimated a net health-sector savings of USD4.95 per person. We also estimated averting 369 disability-adjusted life years (DALYs) per 100,000 population. From a societal perspective, at 10 years *Wolbachia* deployment is expected to have highly favorable

**Conclusions:** Over 10 years, *Wolbachia* is highly beneficial on economic grounds, and almost universally cost saving. That is, *Wolbachia’s* savings in health care costs alone would more than offset deployment costs nationally and in 9 target cities (those with adjusted annual dengue incidence at least 50/100,000 population). In these 9 target cities, *Wolbachia* would generate at least USD3.00 in benefits per dollar invested, giving substantial confidence that *Wolbachia* deployment would be cost-beneficial in Colombia.

## Introduction

Dengue, responsible for dengue fever and dengue hemorrhagic fever, is the most widespread vector-borne virus in the southern hemisphere.[1] Colombia has experienced recent dengue epidemics in 2010, 2013, and 2019.[2]

*Wolbachia* are common bacteria that naturally infect fruit flies and many other insects. Researchers at the World Mosquito Program (WMP) discovered that they could infect *Aedes aegypti* mosquitoes with these bacteria[3] and that dengue, chikungunya and Zika viruses are then less able to replicate within the insects, thereby inhibiting the transmission of these mosquito-borne infections.[4] To use this method for disease control, governments, communities, and international organizations (e.g., the WMP) partner to grow mosquitoes infected with *Wolbachia* in insectaries and then deploy eggs or adult mosquitoes to establish the bacteria in the local mosquito population. *Wolbachia-*infected mosquitoes transmit the bacteria through their eggs to the next generation. This approach is termed the “replacement” strategy, as it tends to replace wild mosquitoes by *Wolbachia*-infected ones. Thus, the establishment of *Wolbachia* becomes a sustainable and often long-term control mechanism at that site. The replacement approach was first applied near Cairns, Australia. Over a decade after initial deployment, mosquitoes there remain infected with the bacteria, supporting the long-term viability of the approach.[5] The replacement approach is being applied in countries in the Americas, Asia, and Oceania.[6]

Under a different approach, the *Wolbachia* suppression strategy, Singapore releases only male *Wolbachia* infected mosquitoes.[3] When these mosquitoes mate with wild mosquitoes, the eggs do not hatch, thereby reducing the number of potentially disease-carrying insects. While experience to date has found this approach efficacious, the need for annual releases makes the suppression approach more costly but potentially economically viable in this high-income country.[7] The remainder of this paper considers only the replacement approach.

A landmark cluster-randomized trial in Yogyakarta, Indonesia found that the replacement strategy reduced all virologically-confirmed symptomatic dengue cases by 77.1% and hospitalized cases by 86.2% under the original protocol analysis.[8] A reanalysis that corrected for the attenuation due to border crossing by humans and mosquitoes raised the estimated efficacy against dengue cases to 82.7%.[9] A subsequent cluster randomized trial is underway in Belo Horizonte, Brazil. A quasi-experimental study from Niterói, Brazil found that *Wolbachia* reduced incidence of dengue by 69%, of chikungunya by 56%, and of Zika by 37%.[10] Research in Rio de Janeiro has shown that the technique is generally robust. Even in neighborhoods where *Wolbachia* coverage was low, such in *favelas* where access was difficult, dengue infections were still reduced by 38% and chikungunya by 10%.[11]

In Colombia, pilot *Wolbachia* releases began in the city of Bello in 2015 and were expanded in 2017 to city-wide deployments throughout Medellín, Itagui and Bello in the Aburrá Valley. An evaluation based on routine disease surveillance data reported reductions in notified dengue incidence of 95% to 97% in the three cities following *Wolbachia* introduction, compared to the prior decade; a parallel case-control study in Medellín also showed significantly lower dengue incidence in *Wolbachia-*treated neighborhoods compared to untreated ones.[12-15] Deployment progressed to Cali, with phased releases since 2020. In May 2023, Cali’s coverage reached 50% and the departmental and municipal governments announced the expansion of *Wolbachia* to Yumbo municipality, 13 km northeast of Cali.[16]

*Wolbachia* is predicted to be a highly cost-effective intervention for controlling mosquito-borne illnesses, especially when released in high-density urban areas. In Indonesia, *Wolbachia* was projected to have a cost-effectiveness ratio in US dollars (USD) of USD1500 per disability-adjusted life year (DALY) averted, offsetting much of the costs to the health system and to society with benefit-cost ratios ranging from 1.35 to 3.40.[17] In Vietnam, another study found the technology similarly cost effective based on the 10-year time horizon, and cost-saving at the 20-year time horizon.[18] A simulation across seven Brazilian cities also found Wolbachia cost-effective across all 7 cities modeled, though not cost saving[19] in 2 of the 7 cities. In Suva, Fiji, a much smaller city, *Wolbachia* was acceptably cost-effective, but in Port Vila, Vanuatu, the relatively small target population and lower population density would not make the approach cost-effective there.[20] A simulation for Thailand suggested that Wolbachia combined with vaccination could be cost-effective.[21]

To inform decision making within Colombia, we modeled the large-scale implementation of the *Wolbachia* replacement strategy for controlling dengue in 11 target Colombian cities. Here we present the resulting cost-effectiveness and benefit-cost analyses.

## Methods

### Framework

The WMP identified 11 target Colombian cities that might be suitable for *Wolbachia* based on population size, population density, and dengue incidence rates, and provided information about each city (Supporting Information S1 Table). Altogether, these cities accounted for a third of Colombia’s reported dengue cases from 2010 through 2019. We conducted economic analyses for each of these target cities. Colombia’s capital and largest city, Bogotá, is virtually free of dengue due to its high altitude, so it was not a target city.

The analyses were done by city, as costs, impacts, and funding decisions lie partly at the municipal level. Our analysis began by estimating the current burden of dengue-related illness in each of these target cities in terms of average annual numbers of cases, health care costs, and loss of health from non-fatal dengue cases. We then estimated the expected gains from *Wolbachia* based on the Yogyakarta cluster randomized trial. Next, we examined the cost of implementing *Wolbachia* based on the WMP’s recent Colombian projects. Finally, we calculated cost-effectiveness and benefit-cost ratios showing the ratio of predicted health care gains to estimated costs by city.

### Parameters

Table 1 provides the necessary national parameters for the economic analysis with monetary amounts in 2020 USD.

**Table 1.**
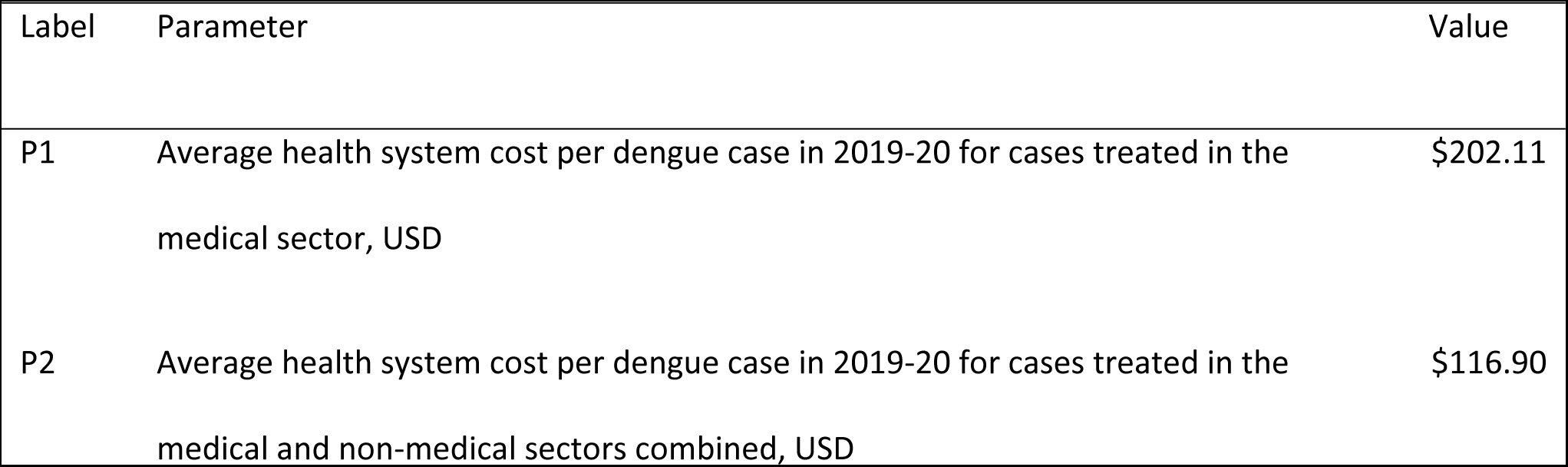
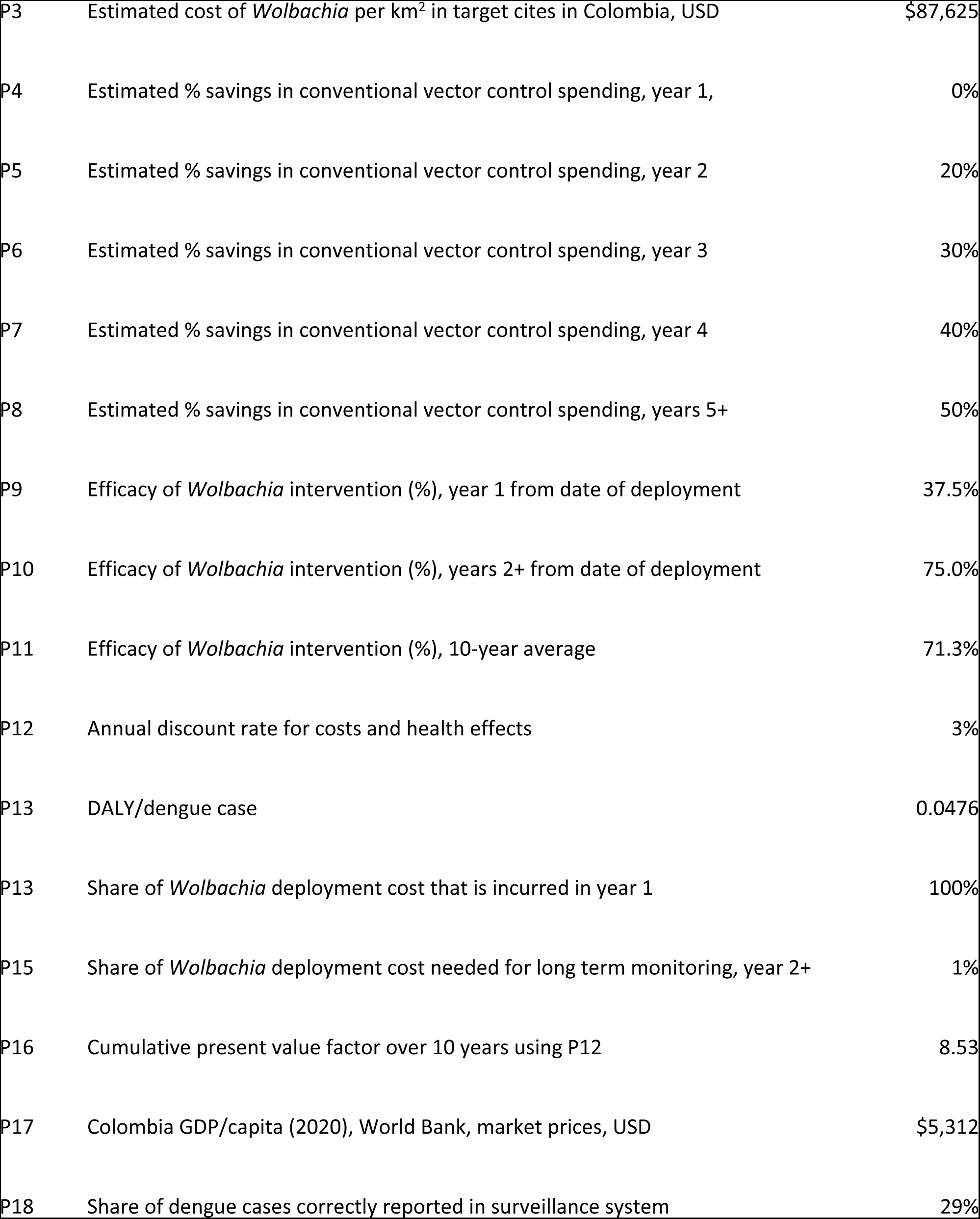
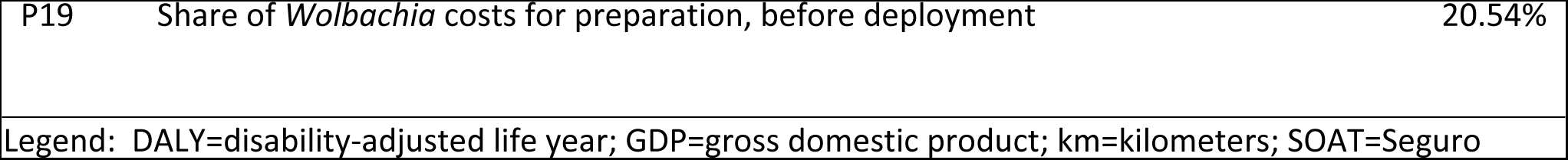
National Parameters.

For P12, we relied on a leading textbook on economic evaluation in health.[22] For P13, the disease burden per case of dengue is the sum of its morbidity and mortality components. The morbidity component was 0.032.[23] The mortality component was calculated first by dividing the average number of deaths due to dengue between the years 2012 through 2018 by the average incidence for these same years. The resulting weighted average case-fatality rate was 6.05 × 10^-4^. Based on an estimated 50 years of remaining life (as young adults are the median age of dengue fatalities) and the widely recommended discount rate of 3%, the discounted remaining life was calculated using the following formula: Discounted remaining life = [1 - (1 + P12)^-50^] /P12 = 25.73. The mortality component was 0.0156 DALYs (i.e. 6.05 × 10^-4^ x 25.73). The overall burden per case was 0.0476 DALYs (i.e., 0.032 + 0.0156).

P16, the cumulative present value factor, was calculated with the Excel present value function (PV) using P12 and a time horizon of 10 years, i.e., PV(P12,10,-1) equals 8.53; P17 is from the World Bank[24]; all other items are based on the authors’ calculations.

### Disease burden of dengue

The disease burden of dengue in a specified geographical area in a year is best conceptualized as the product of its number of dengue cases times the disease burden per case. Global research has found that a substantial share of dengue cases are treated outside the formal health sector, and thus not captured in existing databases.[25] To apply this concept to Colombia, we assessed the breakdown of dengue cases by severity and reporting status. We relied on the expertise of three epidemiologists: Luz Inés Villarreal Salazar (independent consultant in Colombia), Carlos Willian Rincon (University Los Andes), and Maria Patricia Arbelaez Montoya (World Mosquito Program, Colombia). We adjusted for underreporting of the number of dengue cases using an adjustment factor derived from *el Sistema Nacional de Vigilancia en Salud Pública (SIVIGILA)* [the National Public Health Surveillance System] and *Registro Individual de Prestación de Servicios de Salud Municipio de Envigado (RIPS)* [Individual Registry of Provision of Health Services Municipality of Envigado].

To adjust for the fact that routine programs often have fewer resources and less intensive supervision than research trials, we rounded down the per-protocol efficacy from the Indonesian cluster randomized trial.[8] We projected that the *Wolbachia* program in Colombia will result in a 75% reduction in dengue cases once *Wolbachia* is stably established in the mosquito population--the second year of implementation onwards based on projected time for deployment. Projecting a linear increase from zero to complete establishment of *Wolbachia* over the first year of implementation, we estimated a 37.5% reduction in dengue cases overall in the first year.

### Current cost of a dengue episode

The aggregate cost of dengue is the product of the average cost per case times the number of cases. We used two approaches to estimate the cost of a dengue case in Colombia. Under our main approach, the average direct cost of a dengue case treated in the formal health system in 2019 was estimated using the tariffs to pay treatment costs from transit accidents, *Seguro Obligatorio para Accidentes de Tránsito (SOAT*) [Compulsory Insurance for Traffic Accidents]. The SOAT tariffs also serve as reference prices in payment negotiations between insurers and providers. While actual payment rates from other insurers are not publicly available, experts believe that actual payment rates likely average the SOAT rates. Anecdotal reports suggest that in rural areas, where providers are few, providers are paid above the SOAT rates, whereas in urban areas, where providers are numerous, payers can negotiate discounts below the SOAT rates.

We converted the SOAT amounts in Colombian pesos to US dollars at the average exchange rate for the years 2015-2020.[26] For most curative services in the health care system, RIPS provides a national claims system that captures the health care provided to the insured population by diagnostic codes, care provided, and care setting. The data include the number of consultations and procedures used, emergency room visits, and hospitalizations. RIPS categorized dengue cases as classic dengue and severe dengue. For verification we used the *Suficiencia* [Sufficiency] database, which provides service payments for calculating the *Unidad de Pago por Capitación (UPC)* [Capitation Payment Unit] and premium information.[27]

Using the SOAT tariff, we derived the cost per case through stratification by the severity of dengue and calculated a weighted average based on the estimated share of dengue cases by severity. To reflect the fact that a number of non-hemorrhagic (classical) cases were hospitalized, we stratified by severity category instead of treatment setting for consistency among data sources. To report the cost of all types of dengue cases in Colombia from the health system perspective, we adjusted for cases treated outside the health care system. To estimate the economic cost, we incorporated both the cost of cases treated outside the health care system and direct and indirect household expenditures during a dengue episode. We then analyzed the RIPS claims data to derive the average cost of a non-fatal dengue case for the years 2015 through 2020 and reported the average 5-year cost per case based on the severity of dengue, i.e., severe and non-severe dengue. The claims data included the total number and cost of dengue health care services based on the care setting: consultations, procedures, emergencies, and hospitalizations.

To validate our SOAT-based estimate of the healthcare cost per dengue case, we used aggregate data (see Supporting Information S2 Text, Supporting Information S3 Table and Supporting Information S4 Table). [28-33] This aggregate approach, termed macro-costing, used an empirical bed-day equivalent factor of 0.32 of an ambulatory visit compared to an inpatient day[32] and the average cost of a hospitalization and an ambulatory visit. Macro-costing remained a secondary estimate, however, as its applicability rested on the assumption that visits and hospitalizations for dengue consumed the same resources, on average, as visits and hospitalizations for all causes combined.

### Disease burden of dengue per case

Based on the calculation provided for discounted remaining life, the years of life lost and years lost to disability per case are 0.0156 and 0.0320,[23] corresponding to shares of 33% and 67%, respectively. The sum of these two metrics comprised the total disease burden per dengue case of 0.0476 DALYs.

### Cost of *Wolbachia* deployment

To estimate the cost of the *Wolbachia* program in the 11 target cities in Columbia, we started by analyzing the program budget for Cali. The budget covered two programmatic phases, with each phase divided into three stages: prepare, release, and short-term monitoring (STM). The budget covered the administrative and management cost, communication, community engagement, data management, diagnostic, monitoring, mosquito rearing, the release of the *Wolbachia* mosquitoes, surveillance, site start-up, project oversight, and indirect (facilities and administrative) costs.

The preparation stage lasted 12 and 6 months for phases 1 and 2 of the Cali program, respectively; release stages each took 6 months, and the STM stage was 12 months. Initially, the WMP projected that implementation of the *Wolbachia* program would take three years per city. After further review, however, WMP officials and the authors agreed that expansion of the existing program to additional cities in Colombia, and likely in other countries in a scale-up phase, could be achieved with an accelerated timeline and reduced the projected duration.

Based on the shortened timeframes, we reduced the durations of projected staff requirements. We estimated the indirect cost of the *Wolbachia* program at 15% of direct costs. This is the maximum global rate allowed to grantees by the Bill & Melinda Gates Foundation,[28] a major sponsor of *Wolbachia* development. Brandeis researchers also reduced the estimated time needed for preparation, release, and long-term monitoring from 30 months to 15 months. Both adjustments reduced the overall projected per square kilometer (km^2^) cost of the *Wolbachia* program. To estimate the overall cost of the program in the 11 target cities, we made the preceding two adjustments to the budgeted cost of Cali phase 2 deployments to derive an adjusted cost per km^2^ (parameter P5). WMP estimated the projected release area (km^2^) in each target city, including all built-up areas and excluding public spaces, parks, and empty spaces. This area was multiplied by the adjusted cost per km^2^ (parameter P3) to estimate the cost of implementation in the rest of Cali and the 10 other target cities.

Our cost projections generated both best estimates and confidence bounds. The uncertainty reflected alternative estimates of the size of the deployment needed and the cost adjustment attributable to the pause in Cali phase 1 deployment due to the COVID-19 pandemic. The implementation costs occur primarily during the first and second years of release and STM, with an estimated 1% of the initial spending needed annually for long-term monitoring from the second year onward.

### Economic appraisal

We calculated the medical cost offsets from dengue cases averted each year as the cost per symptomatic case times the baseline average number of such cases times the fraction averted in each city year. Although some health economists disagree with discounting future health effects,[34] a leading textbook and Colombia-specific guidelines recommend that future costs and health benefits should be discounted.[22, 35] We calculated the present value of the *Wolbachia* program and all cost offsets in each city over a ten-year time horizon with a discount factor from P12. The vector control offset was calculated through percentagewise cost savings in parameters P4 through P8. The medical cost offset comprises the estimated reduction of cases over the ten-year time horizon. The present value of these offsets was calculated as the annual full-deployment result time the cumulative present value factor for ten years (parameter P16) less an adjustment for the smaller effectiveness in year 1. If we had decided not to discount, then the present value costs would have changed little, but the present value of health impacts would have been substantially higher.

To value the indirect benefits (gains in quality and length of life), we needed to assign an economic value to a year of good health—averting a DALY or gaining a Quality-adjusted Life Year (QALY). This valuation is equivalent to setting a threshold value for determining the cost-effectiveness of a health intervention. In 2001, the World Health Organization’s Macroeconomic Commission on Health recommended thresholds of 1 and 3 times a country’s per capita Gross Domestic Product (GDP) for an intervention to be “very cost-effective” or “cost effective,” respectively.[36] Subsequently, WHO officials recommended finding evidence-based thresholds and incorporating fairness and affordability into the decision process.[37] Economic theory suggests that evidence consider the public’s willingness to pay (WTP) to avert one DALY or gain one QALY.[22]

To apply this concept, we searched PubMed for studies on WTP in Colombia. The one study we found, modeling chemotherapy for lung cancer, did not present an empirical estimate, but simply selected a value of US$17,656, three times Colombia’s then GDP per capita.[38] A wider search, a systematic global review of WTP studies, found a median value for upper-middle income countries (the relevant category for Colombia) of US$5,936, with an interquartile range of US$7,233.[39] However, none of the included studies was conducted in Colombia and upper-middle income countries span a wide range of per capita GDP. However, in 2023, an empirical approach was published for WTP thresholds and applied to 174 countries.[40] It included Colombia. Based on national data rather than survey responses, it calculated national WTPs based on the country’s changes in life expectancy and health expenditures. This approach found that Colombia’s WTP per QALY gained (equivalent to a DALY averted) was 0.75 times its per capita GDP in 2019. An independent commentary noted the many advantages of this approach.[41] Like that in most upper-middle income countries, Colombia’s WTP as a proportion of per capita GDP fell in the range of 0.5 to 1.0. Applying Colombia’s ratio, we valued each DALY in our target year (2020) in Colombia as 0.75 times that year’s per capita GDP. That is, each DALY averted through reduced dengue had an economic value of USD3,984.

We calculated each city’s benefit-cost ratio as its total economic benefits (including the economic value of good health) divided by the cost of the deployment. If this ratio exceeded 1.0, *Wolbachia* was considered a favorable economic investment. The incremental cost-effectiveness ratio (ICER) is the net present value cost of the *Wolbachia* program divided by its present value health gain in DALYs. A positive ICER below USD3,984 (0.75 times Colombia’s per capita GDP[20] of USD5,312) indicates that the intervention is cost-effective. A negative ICER indicates that the replacement strategy is cost saving in that city, i.e., exceptionally cost-effective.

## Results

### Current cost per case of dengue

The epidemiological panel provided the following five categories for the distribution of dengue cases in Colombia by severity and reporting to SIVIGILA: (1) 2% are severe cases and correctly diagnosed and reported to SIVIGILA, (2) 27% are non-severe dengue (including those with and without warning signs) and correctly reported to SIVIGILA, (3) 11% are non-severe dengue, diagnosed by medical providers but not reported to SIVIGILA due to time and administrative barriers, (4) 20% are non-severe dengue cases that are misdiagnosed (e.g., diagnosed as a non-specific viral fever), and (5) 40% are mild and do not interact with the formal healthcare system (i.e. home treatments).

Supporting Information S5 Table presents the average cost of a dengue case by severity and the proportion of dengue cases treated by setting. Only 29% of dengue cases are reported, almost all of which are non-severe dengue. Based on SOAT tariff, we estimated the healthcare cost of care for cases within the medical system as USD406.37 for a severe case (constituting 6.45% of medical cases) and USD188.02 for a non-severe medical dengue case (constituting 93.55% of medical cases). The weighted average healthcare cost per medical case was USD202.11 and USD1.50 for a non-medical dengue case. The societal cost per case averaged USD151.96, comprised of healthcare costs (averaging USD116.90) and indirect costs (averaging USD35.06).

### Economic results in target cities

Table 2 displays the analytic results of *Wolbachia* releases for each target city and the national total (sum of all target cities). Table 3 presents the costs and benefits as rates per person covered and gives final economic results. All benefit cost ratios are favorable or highly favorable, ranging from 1.39 to 8.85.

**Table 2.**
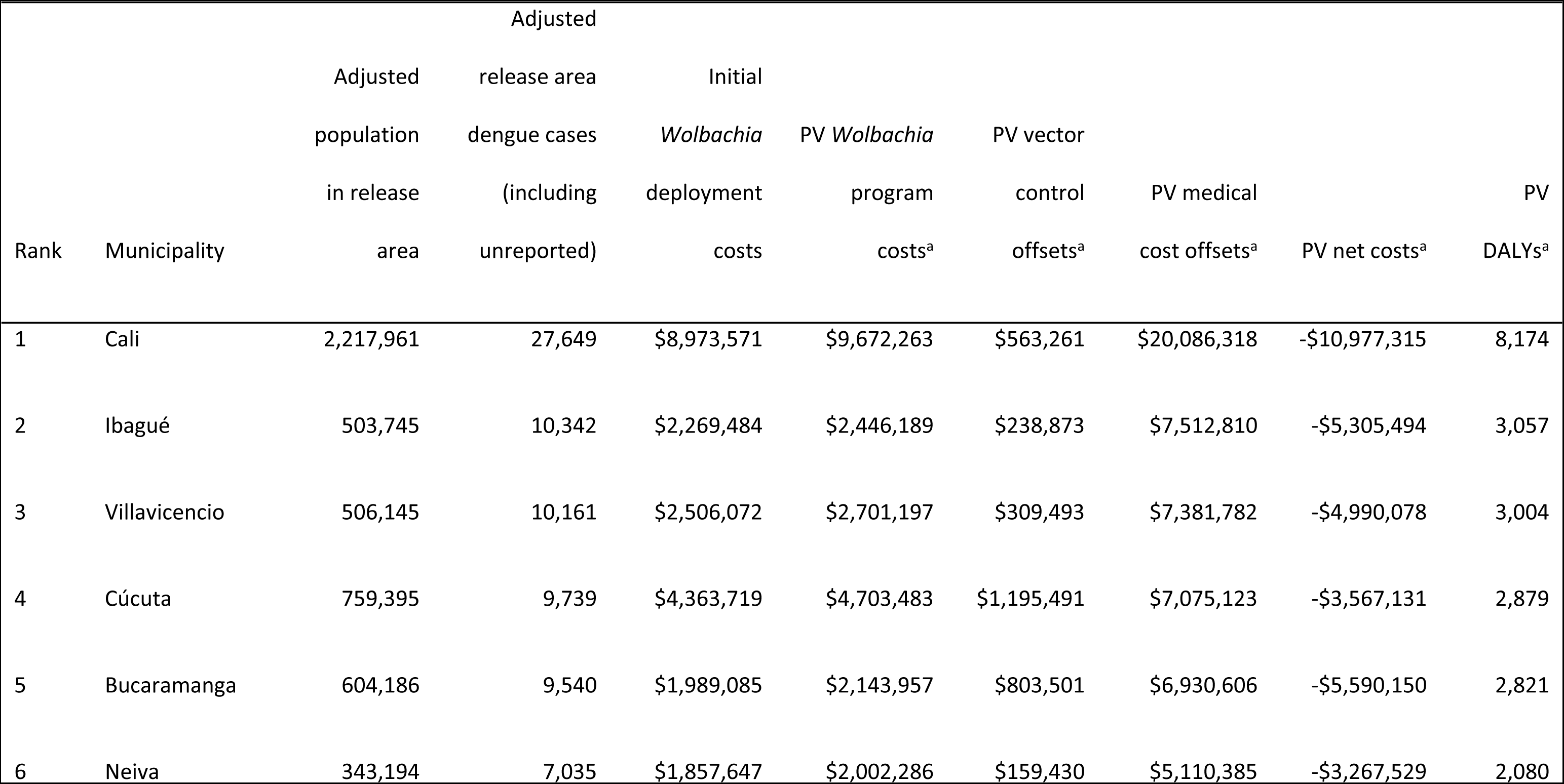
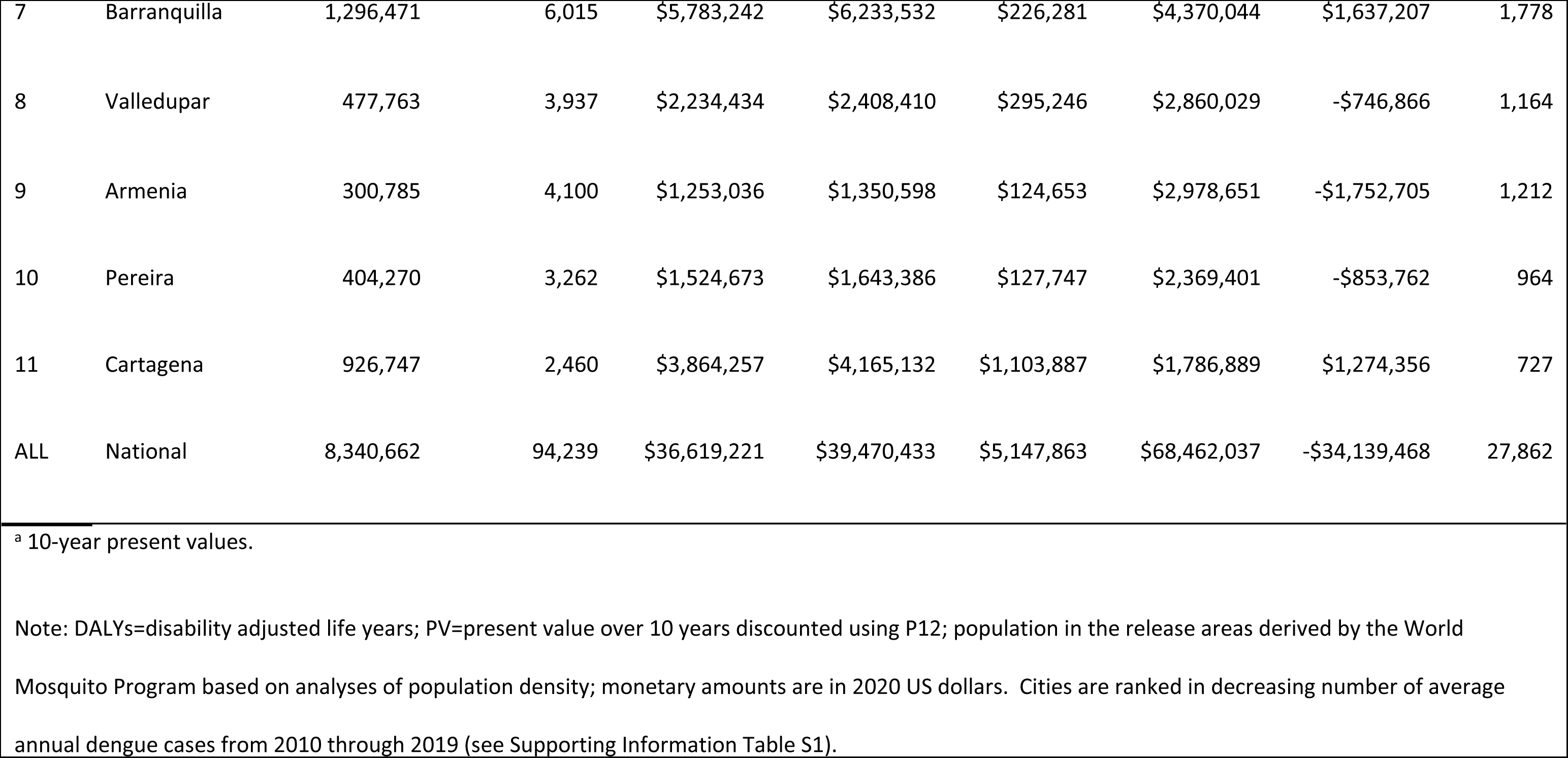
Aggregate costs and DALYs for target cities following the start of *Wolbachia* releases.

**Table 3.**
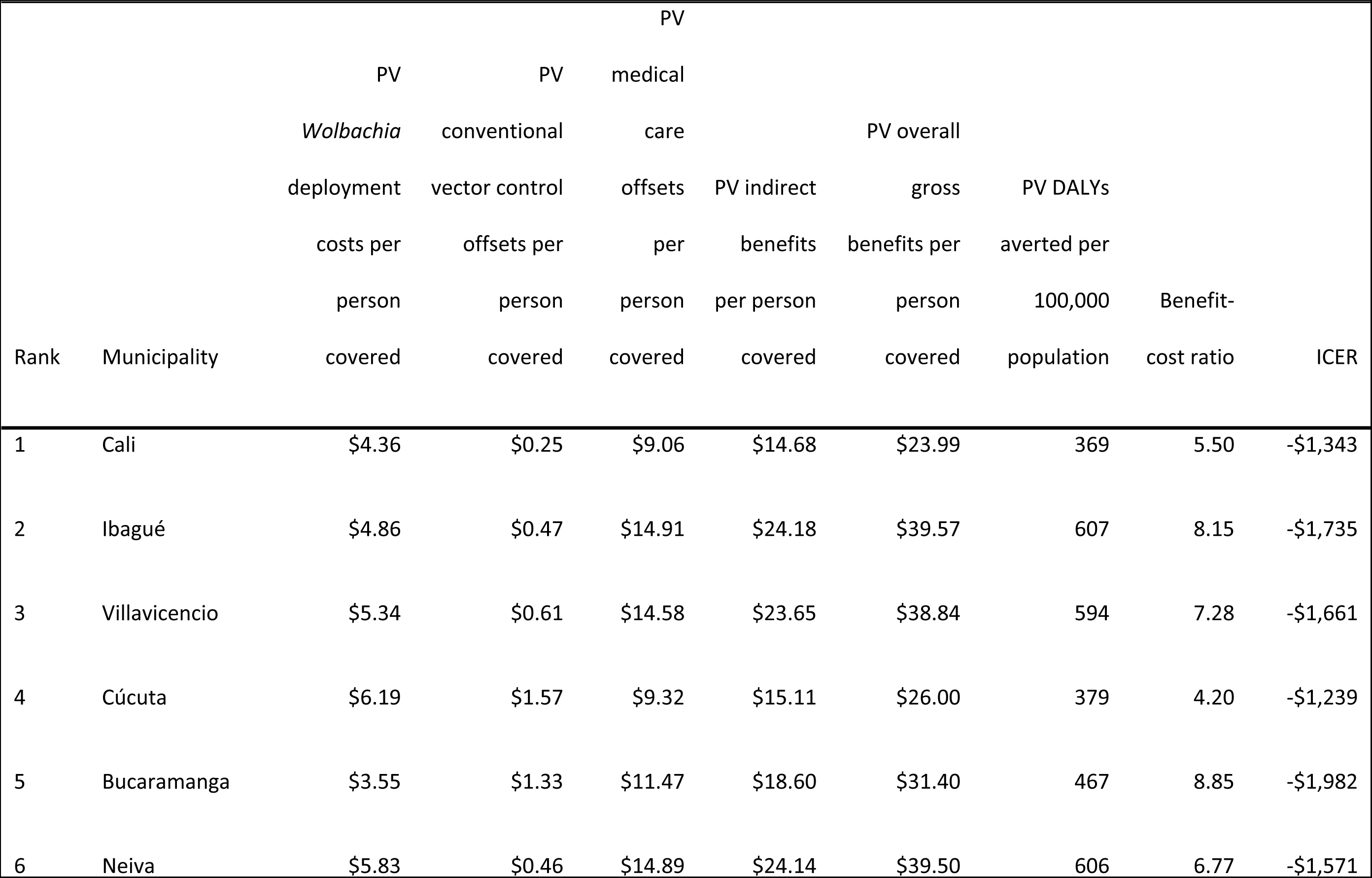
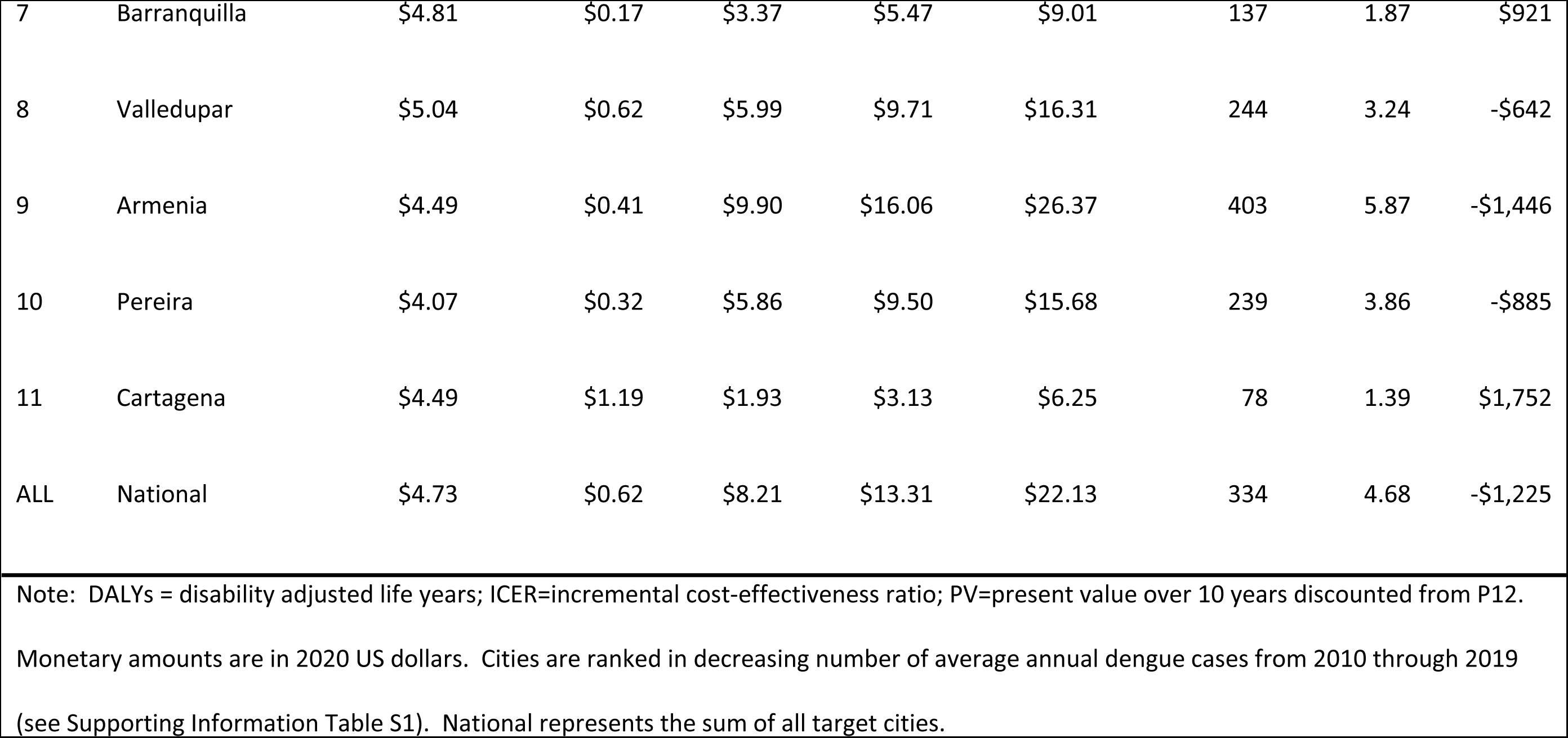
Ratios for target cities following the start of *Wolbachia* releases.

To illustrate our results in greater detail, we have focused on Cali. It was the city with the greatest burden in reported dengue cases. After the Aburrá Valley, where *Wolbachia* had been deployed previously,[12-15] Cali is the one target city in Colombia in which *Wolbachia* is already partly deployed. Fig 1 displays the cumulative projected economic benefits of the *Wolbachia* program in Cali by component and time horizon, where time is the number of completed years since *Wolbachia* deployment. *Wolbachia* is projected to replace some conventional vector control, lower the need for medical care for treating dengue illness, and create economic value of additional healthy years. The overall economic benefits, the sum of these components, grows with increasing time horizons to USD42.97 per person covered with a 20-year horizon. Over this horizon, indirect benefits (the economic value of reduced illness, USD26.27) are the largest component, followed by medical care offsets (USD16.20), with vector control offsets as the smallest benefit (USD0.50),

**Fig 1.**
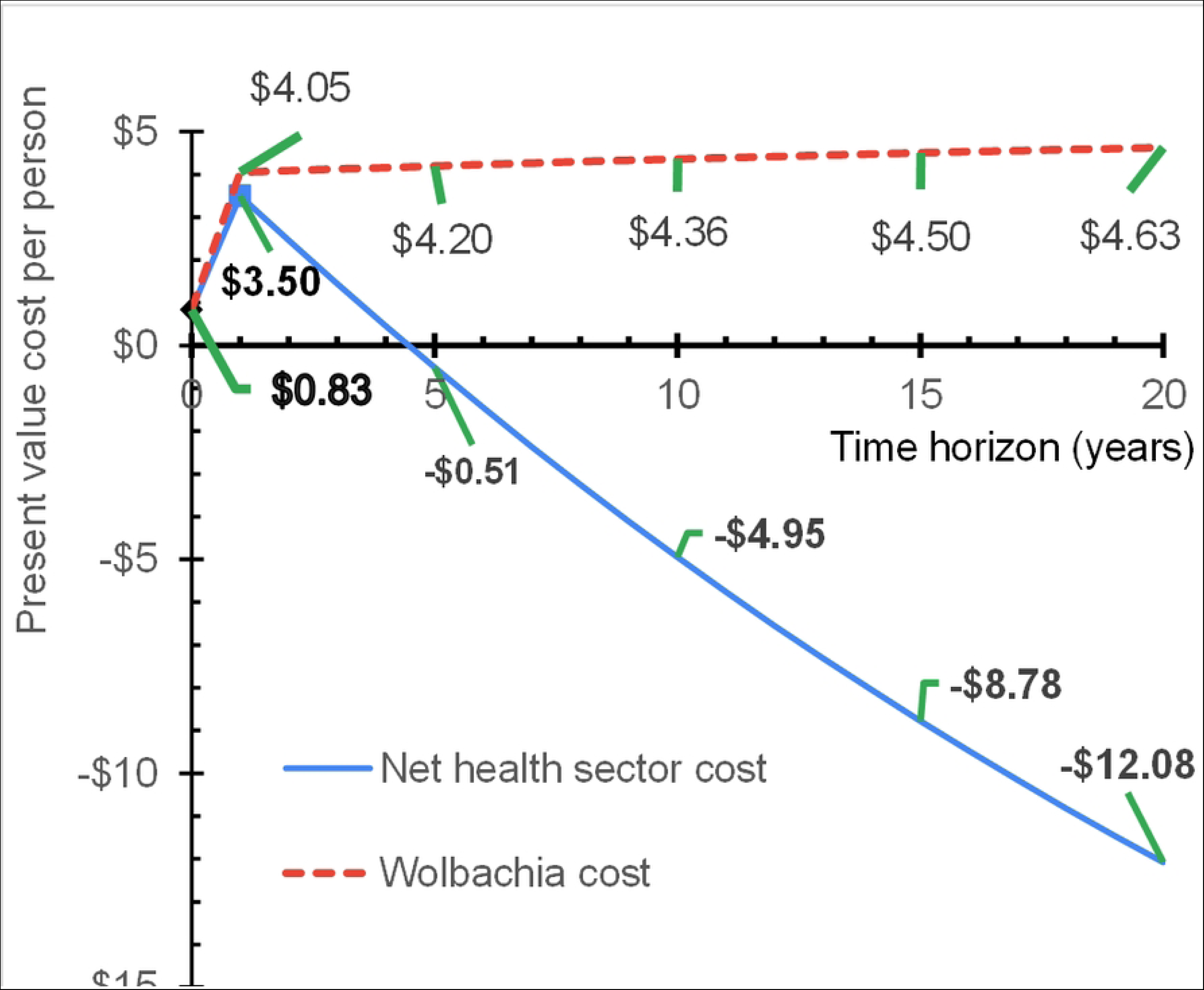
Economic benefits of *Wolbachia* by component and time horizon

The upper (dashed) line in Fig 2 shows the cost per person of implementing the *Wolbachia* program. The cost per person starts in year 0 with 20.54% of initial program costs (USD0.83) for planning and engagement of residents and local leaders. The remainder of initial costs occur in year 1, the year in which city-wide releases would occur, bringing initial program costs to USD4.05. Thereafter, annual monitoring occurs, if needed, costing 1% of the initial costs annually throughout the remainder of the time horizon. Thus, cumulative *Wolbachia* implementation costs per person rise to USD4.20 through 5 years and USD4.63 through 20 years.

**Fig 2.**
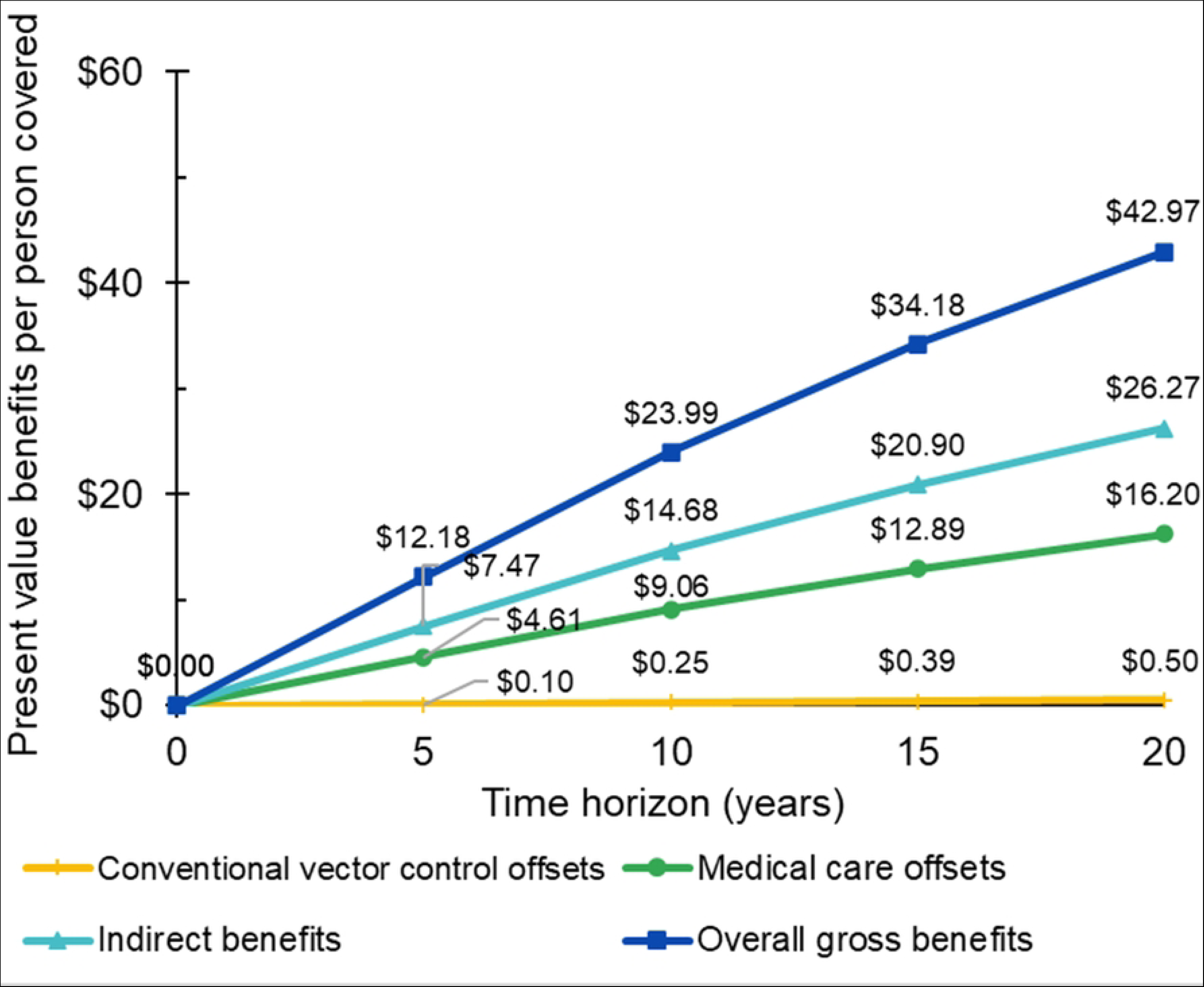
Gross and net healthcare costs of *Wolbachia* by component and time horizon

Whereas longer time horizons generated substantially larger benefits for each of the economic benefits, they added very little to the present value of *Wolbachia* program costs. The lower (solid) line is the net healthcare cost at each time horizon. In year 0, when there are no offsets, it is identical to costs of planning and engagement (USD0.83). In year 1, with some conventional vector control and medical care offsets, net healthcare costs per person covered reached the maximum (USD3.50). In subsequent years, healthcare offsets exceed the additional vector control costs. At 4.3 years, *Wolbachia* becomes cost saving in health care costs. With longer time horizons, the cost savings continue to grow. Net costs per person become substantial negative numbers (-USD4.95 and -USD12.08) with the 10- and 20-year horizons, respectively.

Fig 3 shows the summary outcome measures on health (DALYs averted) and economic impact (benefit-cost ratio) for Cali. Both measures increase with longer time horizons. With a 10-year horizon, the *Wolbachia* program averts 369 DALYs per 100,000 population with a benefit-cost ratio of 5.50. This highly favorable ratio indicates that every dollar invested generates USD5.50 in economic benefits for the city’s residents through better health and averting healthcare costs. With a 20-year horizon, these results become almost twice as favorable, averting 659 DALYs and a benefit-cost ratio of 9.29 to 1. Since the economic benefits from better health and offsets to health care expenses occur approximately uniformly over time, the break-even time horizon at which the overall economic benefits exactly offset the costs is only 1.72 years (21 months) in Cali.

**Fig 3.**
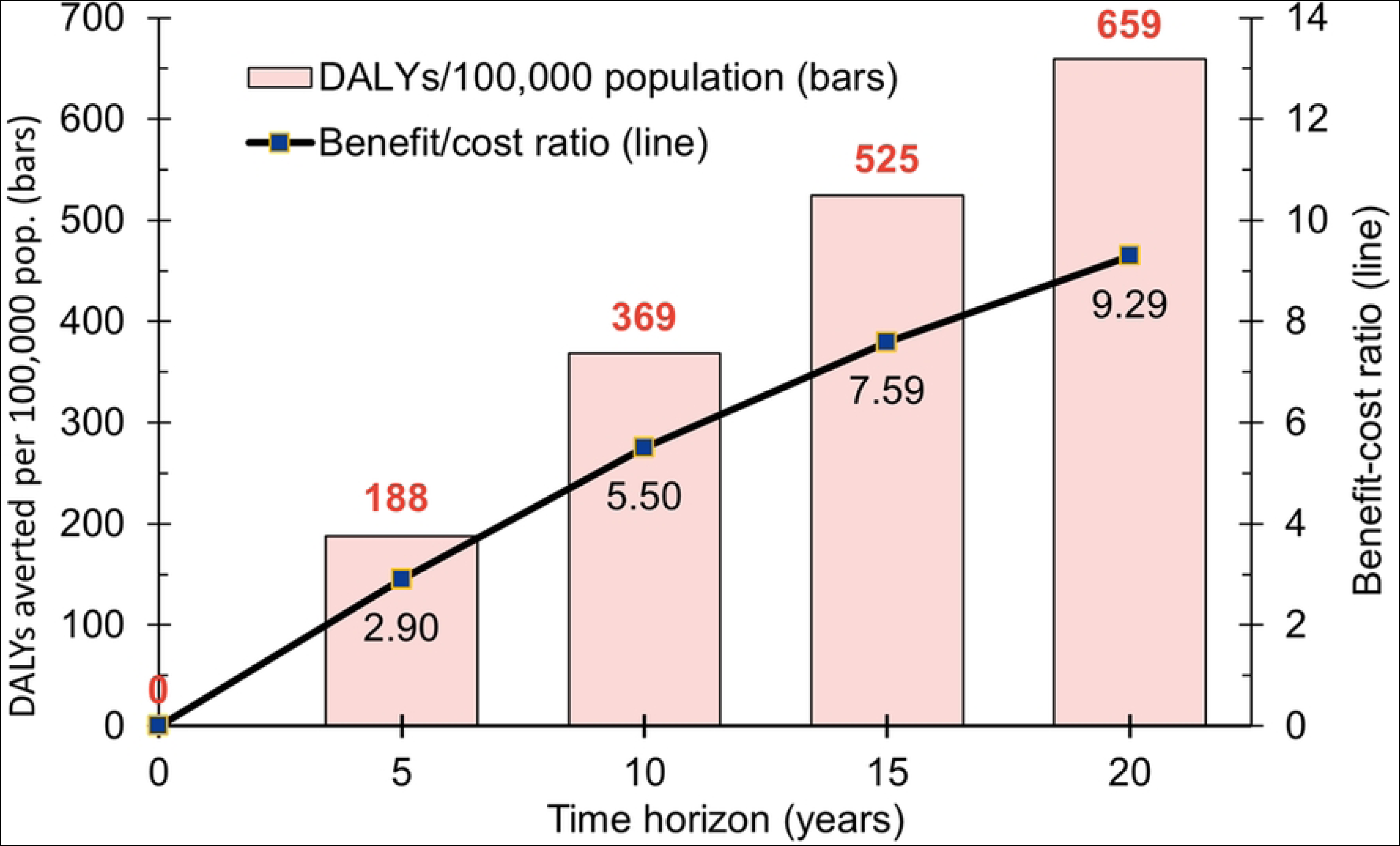
Program impacts (DALYs and benefit-cost ratios) in Cali by time horizon Note: DALYs denote disability adjusted life years

Extending these results nationally, Fig 4 presents the benefit-cost ratios for all target cities based on the 10-year horizon. Panel A displays the cities in decreasing order. Projections for all target cities are favorable, as all the ratios exceed 1.00. Cali is close to the national average. Cartagena is the most marginal in economic terms (ratio 1.39), while Bucaramanga, with a ratio of 8.85, is almost twice as favorable as the national average. Panel B shows a scatter plot of these ratios in relation to population density and average annual dengue incidence.

**Fig 4.**
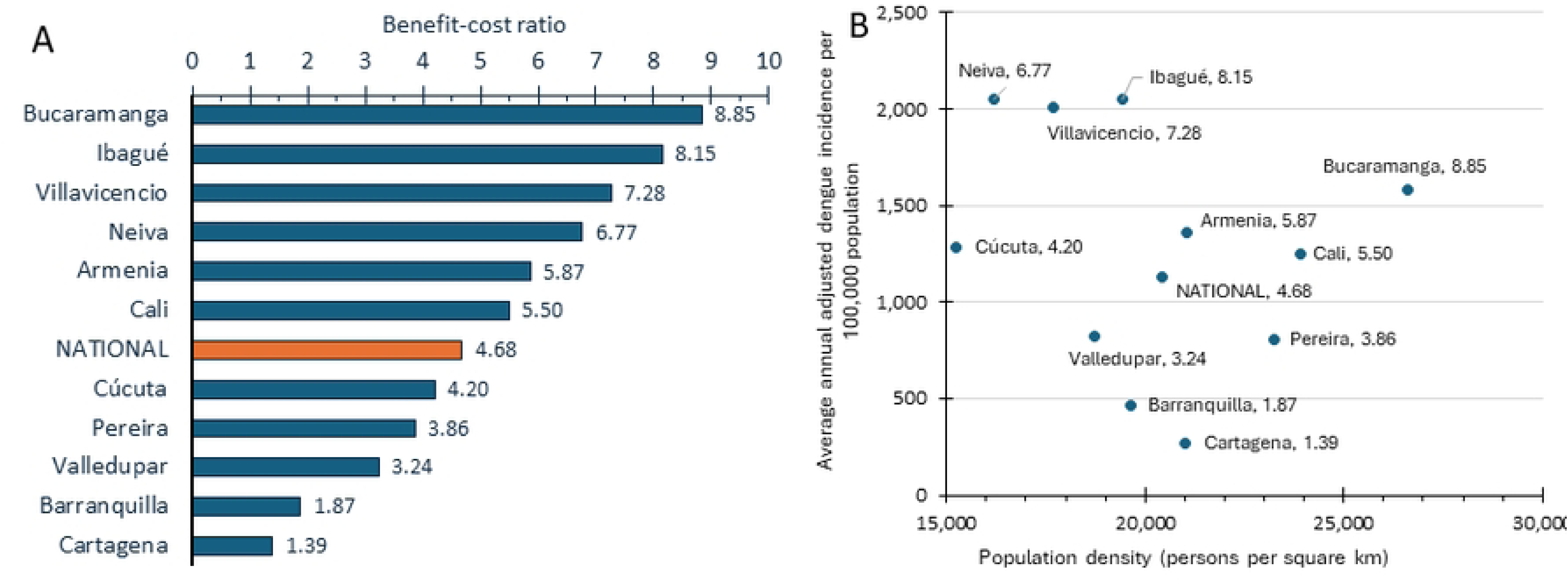
Estimated benefit-cost ratios by city with a 10-year horizon

Higher values of both independent variables tend to be associated with higher (more favorable) benefit-cost ratios. Dengue incidence is the more important factor as it varies 8-fold from the least to the most affected city. Higher population density, which varies by a factor of 1.7, contributes marginally to higher ratios.

## Discussion

Colombia is hyperendemic with dengue.[2] Accounting for cases treated outside the medical system, misdiagnosed, or otherwise not reported, we concluded that the country’s dengue burden is several times greater than official statistics. Our estimates reinforce previous research that Colombia’s dengue burden exceeds the global average.[17, 25]

If implemented with efficacy mirroring the results from the cluster randomized trial,[8] *Wolbachia* will substantially mitigate dengue incidence in the target cities in Colombia. These impacts generate highly favorable benefit-cost ratios by helping to avert healthcare and indirect costs. In over half of the cities, including Cali, the economic benefits exceed USD5.00 for every dollar invested.

*Wolbachia’s* costs mostly occur at the beginning, while the health and economic benefits accrue over time. Therefore, the cost effectiveness and economic benefits of *Wolbachia* increase with longer time horizons. For example, for each dollar invested, the benefit in Cali ranged from USD5.50 at 10 years to USD9.29 at 20 years. Thanks to Colombia’s national health insurance system, the medical care component of these benefits would accrue largely to Colombia’s public sector.

We used a supplemental method to validate the cost per case of dengue through a supplemental calculation. The consistency between our main (SOAT) and supplemental (macro-costing) approaches lent confidence in our results. The difference in cost per case between our main and supplemental approaches (USD117.50 and USD121.61, respectively) was only 3.5%. Because the SOAT approach provided greater detail, it was our preferred choice. We explored performing additional analyses by tier within Colombia’s health system, but inconsistencies precluded doing this reliably with the available data (see Supporting Information S6 Text).

Global experience and models raise a caution that *Wolbachia* may not work in isolated circumstances. As one example, in two nearby sites in Vietnam, *Wolbachia* coverage dropped in one (Tri Nguyan village) but not in the other (Vinh Luong). Researchers speculated that elevated temperature in water storage tanks where mosquitoes bred or an interaction with the built environment may have inhibited *Wolbachia* replication in the ineffective village[42]. As another example, in small-scale releases in Malaysia, *Wolbachia* were not permanently established because the selected strain (wAlbB) may have been less fit than the wild mosquitoes.[42] Modeling studies raise the possibility that dengue viruses could become resistant to *Wolbachia*. Because of the multiple mechanisms by which *Wolbachia* inhibit dengue transmission, however, any such resistance, if any, would likely evolve only slowly.[43] Resistance could be identified by monitoring and possible corrective actions, such as new *Wolbachia* strains.

Our very favorable national benefit-cost ratio of 4.68 indicates that our findings are robust and broadly resistant to such concerns. Our calculations show that the replacement strategy would remain economically viable nationally even if 10-year efficacy declined by as much as 78.6%, calculated as (4.68 – 1)/4.68. With that large a decline costs of USD1.00 would generate benefits of USD4.68 x (100.0% - 78.6%) or USD1.00, meaning that the program would just break even economically.

While our benefit-cost ratios compare *Wolbachia* against no dengue control, policymakers may also wish to consider comparing *Wolbachia* against alternative dengue control strategies. Among the few trials, an alternative vector control strategy based on community-based mobilization (*Camino Verde*) proved to be effective but labor intensive and expensive. Its cost-effectiveness ratios relative to GDP per capita were relatively unfavorable--3.0 in Mexico and 16.9 in Nicaragua.[44] A modeled assessment of screening and vaccination in Colombia with the first licensed dengue vaccine (Sanofi’s Dengvaxia®) gave cost-effectiveness ratios relative to GDP per capita ranging from 0.47 (in areas with 90% dengue seropositivity among 9 year-olds) to 6.72 (with 10% dengue seropositivity).[45] This strategy proved more cost-effective (lower ICER) as the percentage of nine-year old seropositive individuals in the population increased.

The second dengue vaccine, TAK-003 (Takeda’s Qdenga®), was licensed by the European Medicines Agency in 2022 and received pre-approval by the World Health Organization in 2024.[46] Published trial results showed TAK-003 reduced dengue fever cases by 80% and, unlike Dengvaxia®, created no added risk for persons with no prior dengue infection.[46] While preliminary economic models by the manufacturer projected that Qdenga® would be cost saving in Puerto Rico[47] and Thailand[48], we could not find any related peer-reviewed economic studies. Fig 4(B) showed that in the 9 of 11 target municipalities with dengue incidence of at least 500 per 100,000 population, *Wolbachia* also proved cost saving.

As resources for public health interventions are limited, it is informative to compare the cost-effectiveness of *Wolbachia* against that of two other public health preventive interventions in Colombia. First, a year after Colombia had introduced HPV vaccination into its national vaccination program,[49] a cost-effectiveness analysis reported its ICER was greater than three times Colombia’s then GDP per capita, so HPV was not then considered cost-effective.[50] Second, a campaign to encourage COVID-19 vaccination among those at highest risk proved cost-effective [51] by the latest criteria,[40] but not cost saving.

In addition to cost-benefit and cost-effectiveness ratios, policy makers must also consider the affordability of any proposed program. The first year costs of *Wolbachia* deployment ($4.05 per person) represent a notable 0.8% of Colombia’s 2019 per capita health expenditure and might appear too expensive if widely implemented at once. However, the program can become more affordable by phasing deployment across parts of a city over multiple years (as happened in Cali) or sequencing successive cities in different years.

Several limitations should be acknowledged. The number of dengue cases differs between the RIPS and *Suficiencia* databases, pointing to inconsistencies and/or under-reporting. Second, differences in numbers of dengue cases treated among different epidemiological models, macro-costing, RIPS, and SIVIGILA creates uncertainty around the estimated healthcare cost offsets. Finally, our adjustments for underreporting and misdiagnosis are based on our panel’s expert judgment rather than objective information. However, the extremely favorable benefit-cost ratios in 9 or our 11 target cities indicate that *Wolbachia* deployment would still be highly favorable in those cities.

Key strengths also deserve highlighting. First, we believe this is the first economic evaluation of *Wolbachia* in Colombia, building on the empirical record of efficacy and feasibility from the trial in Indonesia[8] and controlled observational studies in the country.[3, 12-15] Second, we used an empirical method for valuing indirect benefits based on the overall economy.[40] As 64% of Colombia’s workers were in the informal sector in 2020 and generally earned less than formal sector workers,[52] this approach is more realistic than applying formal sector wages to indirect costs of all cases, including those not employed or working informally, as was done elsewhere.[19] Third, this study’s number of sites (11) substantially exceeds the numbers in previous economic analyses--3 in Indonesia[17] and 7 in Brazil.[19] These multiple sites provided the insight that not only was *Wolbachia* beneficial overall, but it was especially valuable in cities with high dengue incidence. High population density in the release area was associated with somewhat more favorable outcomes. As a square kilometer with high dengue incidence and high population density is one with substantial dengue burden, deploying *Wolbachia* in such a location will generate substantial economic value. Conversely, areas with relatively low incidence and low density would benefit much less; another control strategy may be preferable.[53]

## Conclusions

In conclusion, *Wolbachia* proved economically beneficial in all 11 target cities and cost saving (paying for itself through treatment costs averted) in the 9 target cities with adjusted incidence of at least 500 per 100,000 population. In the future, policy makers may have a portfolio of options to control dengue. Municipalities with both high incidence of dengue and high population density should strongly consider trying to mobilize the resources for *Wolbachia* deployment. Areas with high dengue incidence but low population density should consider vaccination. To address the uncertainties around each dengue control technique, some experts recommend integrating *Wolbachia*, vaccination, and case management.

## Author Contributions

**Donald S. Shepard**: conceptualization; formal analysis; funding acquisition; methodology; supervision; writing—review & editing. **Samantha R. Lee**: data curation; formal analysis; writing—original draft, review & editing. **Yara A. Halasa-Rappel**: data curation; formal analysis; writing—original draft, review & editing. **Carlos Rincon Perez**: methodology; data curation; formal analysis; writing—review & editing. **Arturo Harker Roa**: methodology; data curation; supervision; writing—review & editing. All authors have read and approved the final version of the manuscript. Donald S. Shepard had full access to all of the data in this study and takes complete responsibility for the integrity of the data and the accuracy of the data analysis.

## Acknowledgments

The authors thank Luz Villarreal Salazar for serving on the study’s epidemiologic panel, Ivan Velez and Patricia Arbelaez Montoya from the WMP Colombia and Reynold Dias and Katherine Anders from the global WMP (Australia) for Wolbachia cost data and valuable comments, and Clare L. Hurley of Brandeis University for editorial assistance.

## Data Availability Statement

Data access may be requested from the respective Colombian government agencies. For RIPS, see https://www.minsalud.gov.co/proteccionsocial/Paginas/rips.aspx. For population, see https://www.dane.gov.co/index.php/estadisticas-por-tema/demografia-y-poblacion/proyecciones-de-poblacion.

## Ethics Statement

This modeling study did not involve any human studies data as it was based entirely on aggregate or publicly available anonymous data. These data could not allow any individual to be identified nor linked with any individual. The research team did not prospectively nor retrospectively recruit human participants nor did the team obtain tissues, data, or samples for the purposes of this study. The research team did not review existing medical records nor archived samples. Therefore, this study was outside the purview of the Committee for Protection of Human Studies in Research so ethical approval was not applicable.

## Transparency Statement

The lead author Donald S. Shepard affirms that this manuscript is an honest, accurate, and transparent account of the study being reported; that no important aspects of the study have been omitted; and that any discrepancies from the study as planned (and, if relevant, registered) have been explained.

## Supporting Information

S1 Table. Input data for target cities

S2 Text: Macro-costing approach

S2 Table. Macro-costing approach to estimate the average cost of an outpatient visit and hospitalization (monetary amounts in 2020 USD)

S4 Table. Health care cost of dengue cases by type of dengue diagnosis and setting using macro-costing (amounts in 2019-2020 USD)

S5 Table. Cost of dengue case by dengue type (Tarifa SOAT for reported), 2019-20 USD S6 Text.

